# Integrating clinical factors and parity-specific models with molecular biomarkers to better predict the risk of preterm birth in asymptomatic women

**DOI:** 10.64898/2026.03.13.26348357

**Authors:** Ashoka Polpitiya, Charles Cox, Heather Butler, Md. Bahadur Badsha, Laura Sommerville, Jay Boniface, George Saade, Paul Kearney

## Abstract

**Background:** Prior spontaneous preterm birth (sPTB) and short cervical length predict the occurrence of sPTB with low sensitivity, highlighting the need for better detectors of at-risk pregnancies. PreTRM® is a validated, biomarker-based sPTB predictor that we aimed to improve in this study by developing models that incorporate parity and key risk factors.

**Methods:** A Model was developed and validated through retrospective analysis of a cohort of singleton pregnancies that resulted in live term or preterm birth (PTB). The Model’s ability to predict sPTB and PTB was assessed and its clinical utility compared to PreTRM.

**Results:** The Model predicted sPTB with 77.1% sensitivity, 74.4% specificity, 21.4% positive predictive value (PPV) and 97.3% negative predictive value (NPV), an improvement over PreTRM’s sensitivity (75.0%) and PPV (14.6%), and higher PPV than short cervix (16.2%). PTB was predicted by the Model with 76.8% sensitivity, 74.6% specificity, 31.6% PPV and 95.5% NPV. The Model predicted a neonatal hospital stay ≥5 days with a significantly higher area under the receiver operating characteristic curve (AUC) than PreTRM associated with PTB (p = 0.001) and sPTB (p = 0.044). The Model also achieved significantly higher sensitivity than PreTRM at predicting a ≥5 day hospital stay associated with PTB (p = 0.009) with improved sensitivity for sPTB, showing overall that the Model performs better than PreTRM in regard to clinical utility.

**Conclusions:** The Model achieved substantially higher performance than standard of care risk predictors, and an improvement in clinical utility over PreTRM, demonstrating the robustness of the Model as a PTB predictor.

## Introduction

Spontaneous preterm birth (sPTB), defined as birth that occurs before 37 weeks of gestational age following spontaneous labor or rupture of the membranes, accounts for an estimated one third of infant deaths in the United States and is a major contributor to neonatal morbidities [1-6]. Various mitigating interventions have been proposed to prevent PTB, including vaginal progesterone, low-dose aspirin, and/or applying a focused care management plan [7-10]. The two best clinical indicators of pregnancies at risk of preterm delivery are prior sPTB and short cervix detected at 18^0/7^ to 22^6/7^ weeks gestation [11-13], yet they have limited impact because the majority of women who experience a sPTB lack premature cervical shortening or a prior sPTB [14-16]. Published findings and calculations made from the literature [7, 14, 17-19] (**Supplemental Figure 1**) indicate that prior sPTB and short cervix identify at-risk women with low sensitivity (6% –11%), highlighting the need for sPTB predictors with better detection rates at similar positive predictive values (PPVs).

As outlined in ACOG Practice Bulletin 234 [20], to be effective, a screening program for identifying women at highest risk for sPTB must have: 1) an adequately high detection rate, 2) appropriately low false positive rate, and 3) an acceptable PPV. We previously validated PreTRM, a serum-based risk predictor indicated for early detection of asymptomatic women at higher risk of sPTB who lack identifiable risk factors including a prior sPTB or short cervix. The PreTRM Test predicts risk based on the ratio of maternal serum insulin-like growth factor binding protein 4 (IGFBP4) to sex hormone binding globulin (SHBG) measured between 18^0/7^ and 20^6/7^ weeks gestation [21, 22]. IGFBP4 is both important for maintaining adequate delivery of nutrients to the fetus and is a marker of placental insufficiency, while SHBG is thought to play a role in the response to placental inflammation via regulation of androgen- and estrogen-driven pathways [23-25]. Placental insufficiency and inflammation are independent causative factors of preterm birth [26-28].

The sensitivity and specificity of the PreTRM Test for predicting sPTB were previously validated at 75.0% and 74.0%, respectively, in a racially diverse, BMI-restricted population with blood drawn between 19^1/7^ and 20^6/7^ weeks (134 –146 days) gestation [21, 22]. In two follow-up clinical utility studies, participants in the screen and treat arm experienced fewer preterm births compared to the control arm. Additionally, the offspring of mothers in the active arm had shorter hospital stays, fewer neonatal intensive care unit (NICU) days, and lower morbidity [29, 30].

Although PreTRM is a validated predictor of sPTB, it does not incorporate parity and only considers pre-pregnancy BMI in its algorithm. Yet, prior pregnancy history is an important predictor of PTB, and risk prediction is impacted by parity. For these reasons, we aimed to develop an improved version of PreTRM that has parity specific sub-models integrated with known clinical risk factors. In addition to developing an improved test, the goals of this study were to 1) compare the improved performance to the standard clinical indicators of prior sPTB and cervical length screening, and 2) compare the performance of the new test to PreTRM with respect to clinical utility. Throughout the paper we refer to this improved version of PreTRM as the ‘Model’.

## Methods

### Cohorts and Study Design

This study was conducted through retrospective analysis of a large, multicenter cohort referred to as PAPR (Proteomic Assessment of Preterm Birth, NCT01371019), which was used to develop the original PreTRM Test [21]. The cohort was restricted to singleton pregnancies, excluded gestations with known or suspected major fetal abnormalities, and included demographic and clinical variables along with serum protein measurements. Data analyzed from the cohort had no missing values.

We developed two models specific to nulliparous and multiparous pregnancies that each used PTB as the clinical outcome. Both models included the log IGFBP4/SHBG ratio (the PreTRM Score) as a predictive factor. Clinical factors were selected for parity-specific model development (**Table 1**) based on guidance in ACOG Practice Bulletin 234 [20]. A nulliparous model that included maternal age and BMI was rejected early in development due to overfitting.

**Table 1.** Clinical factors included in the Model, according to parity.

| **Table 1. Clinical factors included in the Model, according to parity.** |  |  |  |  |  |  |
| --- | --- | --- | --- | --- | --- | --- |
| | Chronic diabetes | BMI ≥ 30 | Maternal age ≥ 35 years | Prior sPTB | Prior PE | Chronic hypertension |
| Nulliparous | ✓ |  |  |  |  |  |
| Multiparous | ✓ | ✓ | ✓ | ✓ | ✓ | ✓ |
| BMI = body mass index, sPTB = spontaneous preterm birth, PE = preeclampsia. |  |  |  |  |  |  |

**Table 2.** Cohort outcomes and demographics.

| **Table 2. Cohort outcomes and demographics.** |  |  |  |
| --- | --- | --- | --- |
| **Clinical Variable** | **Nulliparous** | **Multiparous** | **Combined** |
| **N Total** | 356 | 620 | 976 |
| **N (%) sPTB < 37 Outcome** | 27 (7.6%) | 57 (9.2%) | 84 (8.6%) |
| **N (%) PTB < 37 Outcome** | 49 (13.8%) | 93 (15%) | 142 (14.5%) |
| **N (%) sPTB < 37 & NNLOS ≥ 5 days** | 13 (3.7%) | 23 (3.7%) | 36 (3.7%) |
| **N (%) PTB < 37 & NNLOS ≥ 5 days** | 29 (8.1%) | 36 (5.8%) | 65 (6.7%) |
| **N (%) Chronic Diabetes** | 24 (6.7%) | 31 (5.0%) | 55 (5.6%) |
| **N (%) Chronic Hypertension** | 17 (4.8%) | 44 (7.1%) | 61 (6.3%) |
| **N (%) Prior PE** | N/A | 60 (9.7%) | 60 (6.1%) |
| **N (%) Prior PTB** | N/A | 165 (26.6%) | 165 (16.9%) |
| **NNLOS (mean, median, SD, min, max)** | 4.07, 3, 6.99, 0, 99 | 4.06, 3, 7.93, 0, 88 | 4.06, 3, 7.6, 0, 99 |
| **N (%) NNLOS ≥ 5 days** | 53 (14.9%) | 72 (11.6%) | 125 (12.8%) |
| **Maternal Age (mean, median, SD, min, max)** | 25.29, 24, 5.81, 18, 46 | 28.98, 28, 5.59, 18, 44 | 27.64, 27, 5.94, 18, 46 |
| **N (%) Maternal Age ≥ 35** | 31 (8.7%) | 122 (19.7%) | 153 (15.7%) |
| **BMI (mean, median, SD, min, max)** | 27.45, 25.8, 7.23, 15.2, 58.1 | 29.26, 28.3, 7.59, 15.8, 75.6 | 28.6, 27.3, 7.51, 15.2, 75.6 |
| **N (%) BMI ≥ 30** | 108 (30.3%) | 248 (40.0%) | 356 (36.5%) |
| **N (%) BMI ≥ 21** | 301 (84.6%) | 554 (89.4%) | 855 (87.6%) |
| **N (%) White** | 257 (72.2%) | 450 (72.6%) | 707 (72.4%) |
| **N (%) Black** | 68 (19.1%) | 101 (16.3%) | 169 (17.3%) |
| **N (%) Asian** | 6 (1.7%) | 7 (1.1%) | 13 (1.3%) |
| **N (%) Hispanic** | 98 (27.5%) | 257 (41.5%) | 355 (36.4%) |
| **N (%) Other Race** | 25 (7.0%) | 62 (10.0%) | 87 (8.9%) |
| PE = Preeclampsia; NNLOS = Neonatal Length of (Hospital) Stay; N/A = Not Applicable. |  |  |  |

### Model Development

The PAPR dataset was temporally split into subsets by sample collection date with approximately 50% for training and 50% withheld for validation purposes. A temporal split was chosen to better test the ability for the Model to generalize to a new population. Development of the improved test had three phases. In the first phase, parity-specific models were developed on the first temporal half of the PAPR cohort which included feature selection and bootstrap cross-validation. The second phase validated these parity-specific models on the second temporal half of the cohort to measure performance on an independent set of subjects. Once validated and demonstrating acceptable performance measurements, the parity-specific model coefficients were updated using the validated procedure with the full cohort dataset to ensure that the best possible final model would be deployed for clinical testing [31, 32].

Parity-specific models were fit using ridge regression [33] and the PreTRM Score was standardized to values between 0 and 1 for compatibility with this method. Bootstrap iterations were generated with replacement, each requiring that at least 30% of data be withheld for cross-validation testing. The outcomes in the remaining 70% training portion were upsampled randomly to match the control count and allow fitting methods to prioritize the sensitivity and specificity equally. Average coefficients for the 200 bootstrap iterations were obtained as model parameters to reduce the risk of overfitting. Area under the receiver operating characteristic curves (AUC) were generated for each model category. Models met acceptance criteria if they achieved an AUC > 0.5, a Wilcoxon rank-sum test result with a significance (alpha) of 0.05, and no significant overfitting in the testing sets.

### Model Validation

Models were validated on the 50% withheld temporal split of the cohort. A threshold enrichment and test of significance for the difference in PTB and non-PTB were computed. A secondary analysis was used to test a comprehensive set of models with different outcomes and corresponding thresholds. No correction for multiple testing was applied during the secondary analysis. Once validated, model coefficients were retrained on the full dataset using the validated training methods and mean coefficients from 200 bootstrap iterations were obtained as the model parameters. The combined performance of the nulliparous and multiparous models became the Model.

### Clinical Utility

The Model’s ability to predict a long neonatal hospital stay associated with sPTB and PTB was assessed on the full cohort. For this analysis, cases were defined as a neonatal hospital stay of at least 5 days along with PTB or sPTB respectively and non-cases were defined as any other outcome. The Model performance measured by sensitivity and AUC for these clinical utility outcomes was compared to the PreTRM Test.

### Statistical Analyses

Statistical power for all AUC calculations was determined as previously described [34]. For model training and validation, the ∼50% temporal split of the cohort provided an n = 476 for training and n = 500 for validation (total n = 976) to give a statistical power of 80% for AUC values > 0.59 in the validation. Fisher’s Exact test was used to compute the significance of threshold enrichment with a significance (alpha) of 0.05. Stratification of all PTB and non-PTB was tested using AUC and the Wilcoxon rank-sum test with a significance (alpha) of 0.05. Model calibration was assessed to evaluate the agreement between predicted probabilities and observed outcomes. The degree of miscalibration was quantified using the Estimated Calibration Index (ECI) where ECI with a value of 0 indicating perfect calibration and values below 0.2 representing a high degree of clinical reliability [35]. The statistical significance of differential AUC performance for clinical utility between PreTRM and the Model was computed using the DeLong test with significance (alpha) 0.05. Sensitivities for clinical utility measurements were compared between PreTRM and the Model using the McNemar test, with a significance level (alpha) of 0.05. All 95% confidence intervals (CI) were computed using the Wilson score method. All p-values associated with multiple measurements were adjusted using Bonferroni corrections. All analyses were performed using R version 4.4.2 [36].

## Results

### Study participant demographics

The cohort was comprised of nulliparous (n = 356) and multiparous (n = 620) women with blood drawn between 18^0/7^ and 20^6/7^ weeks gestational age; the intended gestational age for the PreTRM Test. Demographics of study participants grouped into the training and test sets are shown in **Supplemental Tables 1 and 2**.

### Performance of Parity-Specific Models

Performance estimates of the trained nulliparous and multiparous models were evaluated. Both models met acceptance criteria by achieving statistical significance (p < 0.05) and AUCs > 0.5 in both training and test measurements (**Table 3**).

**Table 3.** Model performance and calibration measurements in training/test (bootstrap cross validation) splits of the ∼50% training split of the cohort with PTB as the outcome.

|  |  |  |  |  |  |
| --- | --- | --- | --- | --- | --- |
| Model | Training AUC<br>(95% CI) | Mean AUC of Cross<br>Validation<br>Measurements | Wilcoxon<br>p-value<sup>1</sup> | ECI | R<sup>2</sup> |
| Nulliparous | 0.74 (0.62–0.86) | 0.73 | $< 0.001$ | 0.12 | 0.99 |
| Multiparous | 0.78 (0.71–0.84) | 0.74 | $< 0.001$ | 0.07 | 0.99 |
| <sup>1</sup>P-value showing significance of AUCs ($p < 0.05$) with a Bonferroni correction for multiple testing. . ECI = estimated calibration index. | | | | | |

Model calibration measurements showed little difference between observed and predicted probabilities (data not shown) with estimated calibration index (ECI), falling well within the established threshold for clinical reliability (ECI < 0.2) and a high R^2^ indicating that the model provides accurate estimates. Similar performance and calibration were observed when the models were independently validated on the withheld temporal split of the dataset. The performance of both models combined was validated and also met acceptance criteria (**Table 4**).

**Table 4.** Model performance and calibration measurements in the ∼50% validation subset of the cohort with PTB as the outcome.

| Model | AUC | AUC 95% CI | Wilcoxon p-value<sup>1</sup> | ECI | R<sup>2</sup> |
| --- | --- | --- | --- | --- | --- |
| Nulliparous | 0.71 | 0.60–0.82 | < 0.001 | 0.11 | 0.88 |
| Multiparous | 0.72 | 0.63–0.80 | < 0.001 | 0.09 | 0.72 |
| Combined Parity | 0.70 | 0.64–0.76 | < 0.001 | 0.10 | 0.85 |

<sup>1</sup> P-value showing significance of AUCs ( $p < 0.05$ ) with a Bonferroni correction for multiple testing. ECI = estimated calibration index.

### Performance of Combined Parity Model

After validation, the nulliparous and multiparous model coefficients were retrained on the combined training and validation groups of the cohort to produce the Model. The performance of the Model was then evaluated on two clinical subgroups of the cohort. The first subgroup included the full range of gestational age at blood draw (GABD) (126-146 days) and all BMIs. The second subgroup was restricted to 136-146 days GABD and BMIs ≥21 because the PreTRM Score has been shown to have higher sensitivity under these conditions. The Model produced AUCs ranging from 0.71–0.79, achieving statistical significance for PTB and sPTB in both subgroups (p < 0.05) (**Table 5**).

**Table 5.** Model AUCs for all parities combined for the full cohort.

| **Table 5.** Model AUCs for all parities combined for the full cohort. |  |  |  |  |
| --- | --- | --- | --- | --- |
| **Subgroup** | **Outcome** | **AUC** | **AUC 95% CI** | **Wilcoxon p-value<sup>1</sup>** |
| Full GABD range, all BMIs | sPTB | 0.71 | 0.65–0.77 | < 0.001 |
|  | PTB | 0.73 | 0.69–0.78 | < 0.001 |
| Restricted GABD range and BMI | sPTB | 0.77 | 0.70–0.85 | < 0.001 |
|  | PTB | 0.79 | 0.72–0.85 | < 0.001 |
| <sup>1</sup>P-value showing significance of AUCs ($p < 0.05$) with a Bonferroni correction for multiple testing. | | | | |

We next evaluated the clinical performance of the Model. In the restricted subgroup the Model detected sPTB with 77.1% sensitivity (95% CI, 59.4–89.0), 74.4% specificity (95% CI, 69.7–78.6), 21.4% PPV, and 97.3% NPV. In this same population it detected PTB with 76.8% sensitivity (95% CI, 63.3–86.6), 74.6% specificity (95% CI, 69.7–78.9), 31.6% PPV, and 95.5% NPV (**Table 6**). The Model demonstrated similar specificities and lower sensitivities in the unrestricted subgroup. The number needed to screen (NNS) to detect either a single PTB or a single sPTB were both higher in the unrestricted subgroup relative to the restricted subgroup.

**Table 6.** Model performance on the full cohort for all parities combined. The 95% confidence intervals were computed using binomial proportions.

|  |  |  |  |  |  |  |  |  |  |
| --- | --- | --- | --- | --- | --- | --- | --- | --- | --- |
| **Subgroup** | **Outcome** | **Fisher's p-value<sup>1</sup>** | **Sensitivity (%)**<br>**(95% CI)** | **Specificity (%)**<br>**(95% CI)** | **PPV (%)**<br>**(95% CI)** | **NPV (%)**<br>**(95% CI)** | **LR+** | **LR-** | **NNS**<br>**(95% CI)** |
| Full GABD range, all BMIs | sPTB | < 0.001 | 61.9 (50.6–72.1) | 76.7 (73.7–79.4) | 20.0 (16.9–23.5) | 95.5 (94.2–96.6) | 2.65<br>(2.16–3.26) | 0.5<br>(0.38–0.65) | 19<br>(16–23) |
|  | PTB | < 0.001 | 63.4 (54.8–71.2) | 76.7 (73.7–79.5) | 31.7 (28.0–35.6) | 92.5 (90.8–93.9) | 2.72<br>(2.29–3.25) | 0.48<br>(0.38–0.59) | 11<br>(10–13) |
| Restricted GABD range and BMI | sPTB | < 0.001 | 77.1 (59.4–89.0) | 74.4 (69.7–78.6) | 21.4 (17.6–25.9) | 97.3 (95.1–98.5) | 3.02<br>(2.35–3.86) | 0.31<br>(0.17–0.57) | 16<br>(14–20) |
|  | PTB | < 0.001 | 76.8 (63.3–86.6) | 74.6 (69.7–78.9) | 31.6 (26.9–36.7) | 95.5 (92.9–97.1) | 3.02<br>(2.41–3.79) | 0.31<br>(0.19–0.5) | 10 (9–12) |
| <sup>1</sup> P-values showing the significance of the chosen threshold ($p < 0.05$) with a Bonferroni correction for multiple testing. PPV = positive predictive value; NPV = negative predictive value; NNS = number needed to screen to detect one (s)PTB. | | | | | | | | | |

Importantly, the Model demonstrated similar PPV (21.4%) to the currently used risk predictors, prior sPTB (22.5%) and cervical length screening (16.2%), while having a far superior sensitivity (77.1% vs 8% and 11%) (**Table 7**).

**Table 7.**
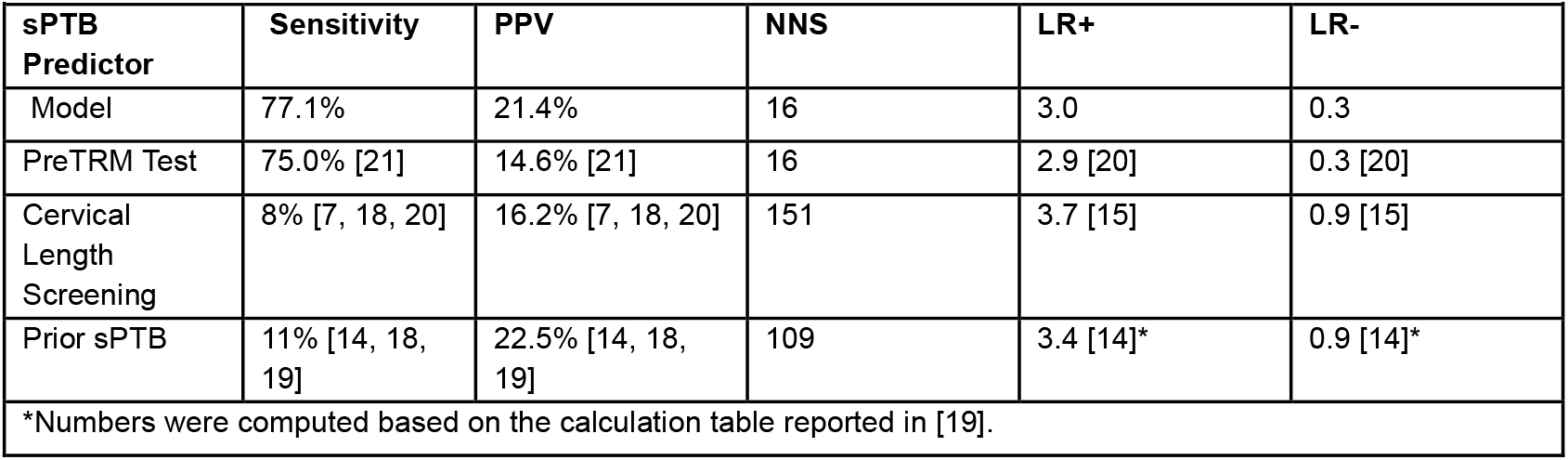
Comparisons between the Model, the PreTRM Test, and today’s clinical predictors of spontaneous preterm birth (sPTB) using key performance metrics as specified by ACOG (Bulletin 234). NNS was calculated using the PAPR sPTB prevalence.

### Assessment of Clinical Utility

We next compared the performance of the Model to PreTRM at predicting if a PTB or sPTB was associated with a neonatal length of stay (LOS) in the hospital lasting 5 days or more. This assessment is important as it demonstrates whether the Model is as good or better than PreTRM at selecting pregnancies that could benefit from intervention. PreTRM and the Model both demonstrated statistically significant AUC values for PTB predictions, and the Model was significant for sPTB when corrected for multiple comparisons (**Table 8**). The Model achieved significantly higher AUC and sensitivity than PreTRM (p = 0.001, and p = 0.009 respectively) at predicting LOS > 5 days associated with PTB. Relative to PreTRM the Model demonstrated a higher AUC and sensitivity at predicting LOS associated with sPTB, which was significant for AUC but did not reach statistical significance for sensitivity (p = 0.044, and p = 0.263 respectively). Overall, these findings show that the clinical utility demonstrated by the Model is an improvement over that of PreTRM.

**Table 8.** Performance comparisons between the Model and the PreTRM Test in predicting a clinical outcome. Note that LOS≥5 refers to a neonatal hospital stay of 5 or more days.

| Metric | Model |  |  |  | PreTRM Test |  |  |  | DeLong<br>p-value<sup>2</sup> | McNemar<br>p-value<sup>3</sup> |
| --- | --- | --- | --- | --- | --- | --- | --- | --- | --- | --- |
|  | AUC<br>(95%<br>CI) | Wilcoxon<br>p-value<sup>1</sup> | Sensitivity<br>(95% CI) | Specificity<br>(95% CI) | AUC<br>(95%<br>CI) | Wilcoxon<br>p-value<sup>1</sup> | Sensitivity<br>(95% CI) | Specificity<br>(95% CI) |  |  |
| LOS≥5 +<br>sPTB | 0.74<br>(0.66 -<br>0.82) | <0.001 | 69.4 (51.7<br>- 83.1) | 75.0 (72.1<br>- 77.7) | 0.61<br>(0.50 -<br>0.72) | 0.054 | 55.6 (38.3<br>- 71.7) | 67.9 (64.8<br>- 70.8) | 0.044 | 0.263 |
| LOS≥5 +<br>PTB | 0.76<br>(0.71 -<br>0.82) | <0.001 | 72.3 (59.6<br>- 82.3) | 74.0 (71.0<br>- 76.8) | 0.62<br>(0.54 -<br>0.70) | 0.002 | 52.3 (39.6<br>- 64.7) | 68.4 (65.2<br>- 71.4) | <0.001 | 0.009 |
| <sup>1</sup>P-value showing significance of AUC (p < 0.05) with a Bonferroni correcting for multiple testing. <sup>2</sup>P-values comparing AUCs (p < 0.05) between the Model and PreTRM with a Bonferroni correction for multiple testing. <sup>3</sup>P-values comparing Sensitivities (p < 0.05) between the Model and PreTRM with Bonferroni correction for multiple testing. |  |  |  |  |  |  |  |  |  |  |

## Discussion

Survival and long-term health outcomes of premature infants could be improved by extending the length of gestation. Tools that can reliably predict pregnancies at risk of preterm birth and benefit from treatment strategies shown to impact clinical outcomes are a significant need [30]. The goal of this study was to improve the performance of PreTRM by developing a Model that integrated parity and other clinical factors with the log IGFBP4/SHBG ratio (PreTRM Score) then validating it for the prediction of sPTB and PTB. The resulting Model predicted sPTB with 77.1% sensitivity, 74.4% specificity, 21.4% PPV and 97.3% NPV. For the prediction of PTB the Model achieved 76.8% sensitivity, 74.6% specificity, 31.6% PPV and 95.5% NPV. Overall, the Model outperformed PreTRM in sensitivity and PPV, important metrics for tests designed to screen patients for intervention. Importantly, in contrast to PreTRM, the Model reported performance for a wider BMI range, and both training and validation included a larger number of subjects (n = 476 and n = 500). In a direct comparison, the Model significantly outperformed PreTRM in predicting a long neonatal hospital stay associated with PTB. The Model also predicted long hospital stay associated with sPTB with a higher AUC and sensitivity than PreTRM, which was significant for AUC but did not reach statistical significance for sensitivity. Overall, the Model’s performance was an improvement over PreTRM at detecting pregnancies susceptible to sPTB and superior at detecting pregnancies at risk of PTB.

Another significant finding of this study was the Model’s performance relative to the performances of short cervix and prior sPTB for screening pregnancies at risk of sPTB. The Model predicted sPTB with a higher PPV (21.4%) than short cervix (16.4%) and a similar PPV to prior sPTB (22.5%) while having a substantially higher sensitivity (77%) than those calculated for both short cervix (8%) and prior sPTB (11%). This improvement in PPV is particularly noteworthy because it brings the test in line with stated performance expectations and endorsed screening approaches [21, 30, 38].

Strengths of this study include the use of a larger cohort for test development and assessment as well as assessment across diverse populations. Although this study was limited by its retrospective design, our findings represent another important step towards improving risk stratifiers to help identify and prevent preterm deliveries and subsequent complications.

## Data Availability

All data produced in the present work are contained in the manuscript

