## Supplemental Figure 1 for "Integrating clinical factors and parity-specific models with molecular biomarkers to better predict the risk of preterm birth in asymptomatic women"

### Supplemental Materials

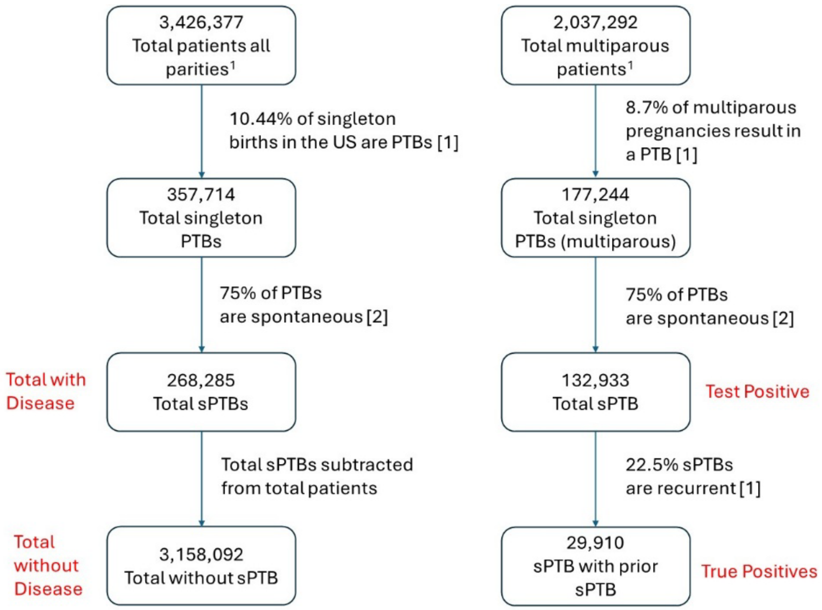

| Definitions | Calculations and Values |
|---|---|
| True Positive- sPTB with prior sPTB | 29,910 |
| False Negative- sPTB without prior sPTB | (268,285 – 29,910) = 238,375 |
| False Positive- No PTB with prior sPTB | (132,933 – 29,910) = 103,023 |
| True Negative- No PTB, no prior sPTB | (3,158,092 – 103,023) = 3,055,069 |
| **Performance Metric** | **Value** |
| Sensitivity | 11.1% |
| Specificity | 96.7% |
| PPV | 22.5% |
| NPV | 92.8% |

**Supplemental Figure 1. Performance metrics of prior sPTB as a predictor of at-risk pregnancies.** (**Left**) Flow chart showing how clinical values were calculated. (**Right**) Chart showing performance metrics and calculations.

[1] Total patients are defined as women who received prenatal care within the first 4 months of pregnancy, required by Petrini et al., for treatment eligibility.

[1] Petrini JR, Callaghan WM, Klebanoff M, Green NS, Lackritz EM, Howse JL, et al. Estimated effect of 17 alpha-hydroxyprogesterone caproate on preterm birth in the United States. *Obstet Gynecol.* 2005;105(2):267-72.

[2] Martin JA, Hamilton BE, Sutton PD, Ventura SJ, Menacker F, Munson ML. Births: Final Data for 2002. *Natl Vital Stat Rep.* 2003; 52(10):1-113
