## Supplemental Table 1 for "Integrating clinical factors and parity-specific models with molecular biomarkers to better predict the risk of preterm birth in asymptomatic women"

| **Supplemental Table 1.** Demographics of PAPR training set. | | |
|---|---|---|
| **Clinical Variable** | **Nulliparous** | **Multiparous** |
| **N Total** | 155 | 321 |
| **N (%) sPTB < 37 Outcome** | 13 (8.4%) | 27 (8.4%) |
| **N (%) PTB < 37 Outcome** | 19 (12.3%) | 53 (16.5%) |
| **N (%) Chronic Diabetes** | 11 (7.1%) | 18 (5.6%) |
| **N (%) Chronic Hypertension** | 5 (3.23%) | 27 (8.41%) |
| **N (%) Prior PE** | N/A | 32 (10.0%) |
| **N (%) Prior sPTB** | N/A | 86 (26.8%) |
| **NNLOS (mean, median, SD, min, max)** | 3.06, 2, 3.52, 0, 32 | 3.82, 3, 5.94, 0, 71 |
| **N (%) NNLOS ≥ 5 days** | 13 (8.4%) | 42 (13.1%) |
| **Maternal Age (mean, median, SD, min, max)** | 24.67, 23, 5.43, 18, 40 | 29.13, 29, 5.48, 18, 43 |
| **N (%) Maternal Age ≥ 30** | 34 (21.9%) | 142 (44.2%) |
| **N (%) Maternal Age ≥ 35** | 11 (7.1%) | 60 (18.7%) |
| **BMI (mean, median, SD, min, max)** | 26.89, 25.2, 6.88, 15.2, 51.4 | 29.22, 28.3, 7.57, 17, 75.6 |
| **N (%) BMI ≥ 30** | 46 (29.7%) | 129 (40.2%) |
| **N (%) BMI ≥ 21** | 127 (81.9%) | 284 (88.5%) |
| **N (%) White** | 113 (72.9%) | 236 (73.5%) |
| **N (%) Black** | 29 (18.7%) | 47 (14.6%) |
| **N (%) Asian** | 1 (0.6%) | 4 (1.2%) |
| **N (%) Hispanic** | 51 (32.9%) | 130 (40.5%) |
| **N (%) Other Race** | 12 (7.7%) | 34 (10.6%) |
