## Supplemental Table 2 for "Integrating clinical factors and parity-specific models with molecular biomarkers to better predict the risk of preterm birth in asymptomatic women"

**Supplemental Table 2.** Demographics of PAPR test (validation) set.

| Clinical Variable | Nulliparous | Multiparous |
|---|---|---|
| **N Total** | 201 | 299 |
| **N (%) sPTB < 37 Outcome** | 14 (6.97%) | 30 (10.03%) |
| **N (%) PTB < 37 Outcome** | 30 (14.93%) | 40 (13.38%) |
| **N (%) Chronic Diabetes** | 13 (6.47%) | 13 (4.35%) |
| **N (%) Chronic Hypertension** | 12 (5.97%) | 17 (5.69%) |
| **N (%) Prior PE** | N/A | 28 (9.36%) |
| **N (%) Prior sPTB** | N/A | 70 (23.41%) |
| **NNLOS (mean, median, SD, min, max)** | 4.85, 3, 8.73, 0, 99 | 4.31, 3, 9.61, 0, 88 |
| **N (%) NNLOS ≥ 5 days** | 40 (19.9%) | 30 (10.03%) |
| **Maternal Age (mean, median, SD, min, max)** | 25.77, 24, 6.05, 18, 46 | 28.82, 28, 5.7, 18, 44 |
| **N (%) Maternal Age ≥ 30** | 55 (27.36%) | 127 (42.47%) |
| **N (%) Maternal Age ≥ 35** | 20 (9.95%) | 62 (20.74%) |
| **BMI (mean, median, SD, min, max)** | 27.89, 26, 7.47, 16.6, 58.1 | 29.31, 28.2, 7.62, 15.8, 61.4 |
| **N (%) BMI ≥ 30** | 62 (30.85%) | 119 (39.8%) |
| **N (%) BMI ≥ 21** | 174 (86.57%) | 270 (90.3%) |
| **N (%) White** | 144 (71.64%) | 214 (71.57%) |
| **N (%) Black** | 39 (19.4%) | 54 (18.06%) |
| **N (%) Asian** | 5 (2.49%) | 3 (1%) |
| **N (%) Hispanic** | 47 (23.38%) | 127 (42.47%) |
| **N (%) Other Race** | 13 (6.47%) | 28 (9.36%) |
